# Safety of Hydroxychloroquine among Outpatient Clinical Trial Participants for COVID-19

**DOI:** 10.1101/2020.07.16.20155531

**Authors:** Sarah M Lofgren, Melanie R Nicol, Ananta S Bangdiwala, Katelyn A Pastick, Elizabeth C Okafor, Caleb P Skipper, Matthew F Pullen, Nicole W Engen, Mahsa Abassi, Darlisha A Williams, Alanna A Nascene, Margaret L Axelrod, Sylvain A Lother, Lauren J MacKenzie, Glen Drobot, Nicole Marten, Matthew P Cheng, Ryan Zarychanski, Ilan S Schwartz, Michael Silverman, Zain Chagla, Lauren E Kelley, Emily G McDonald, Todd C Lee, Katherine H Hullsiek, David R. Boulware, Radha Rajasingham

## Abstract

**Introduction:** Use of hydroxychloroquine in hospitalized patients with COVID-19, especially in combination with azithromycin, has raised safety concerns. Here, we report safety data from three outpatient randomized clinical trials.

**Methods:** We conducted three randomized, double-blind, placebo-controlled trials investigating hydroxychloroquine as pre-exposure prophylaxis, post-exposure prophylaxis and early treatment for COVID-19. We excluded individuals with contraindications to hydroxychloroquine. We collected side effects and serious adverse events. We report descriptive analyses of our findings.

**Results:** We enrolled 2,795 participants. The median age of research participants was 40 (IQR 34-49) years, and 59% (1633/2767) reported no chronic medical conditions. Overall 2,324 (84%) participants reported side effect data, and 638 (27%) reported at least one medication side effect. Side effects were reported in 29% with daily, 36% with twice weekly, 31% with once weekly hydroxychloroquine compared to 19% with placebo. The most common side effects were upset stomach or nausea (25% with daily, 18% with twice weekly, 16% with weekly, vs. 10% for placebo), followed by diarrhea, vomiting, or abdominal pain (23% for daily, 16% twice weekly, 12% weekly, vs. 6% for placebo). Two individuals were hospitalized for atrial arrhythmias, one on placebo and one on twice weekly hydroxychloroquine. No sudden deaths occurred.

**Conclusion:** Data from three outpatient COVID-19 trials demonstrated that gastrointestinal side effects were common but mild with the use of hydroxychloroquine, while serious side effects were rare. No deaths occurred related to hydroxychloroquine. Randomized clinical trials can safely investigate whether hydroxychloroquine is efficacious for COVID-19.

**Short Summary:** Data from three randomized clinical trials using hydroxychloroquine for the prevention and treatment of COVID-19 did not suggest significant safety concerns. Gastrointestinal side effects were common but arrhythmias were rare. There were no sudden deaths in any trial.

## Introduction

Hydroxychloroquine has *in vitro* antiviral activity against *Severe Acute Respiratory Syndrome-related coronavirus 2* (SARS-CoV-2).[1, 2] While *in vitro* data suggest that both chloroquine and hydroxychloroquine have activity against SARS-CoV-2 replication, the latter is generally considered less toxic and better tolerated. However, clinical data to date demonstrate no conclusive efficacy of hydroxychloroquine for the treatment or prevention of COVID-19.[3-5] Both chloroquine and hydroxychloroquine impede SARS-CoV-2 replication.[2] Several randomized placebo-controlled clinical trials are underway to evaluate hydroxychloroquine’s safety and efficacy in the prevention and treatment of COVID-19 in both inpatient and outpatient populations.[6]

In late March 2020, hydroxychloroquine had substantial positive coverage in the media. However, the tide rapidly turned due to concerted efforts to inform physicians and patients about the potential risks of taking the drug outside of clinical trial settings.[7] Several inpatient treatment studies then went on to show increased cardiac side effects with hydroxychloroquine and azithromycin.[8, 9] On April 24th, 2020, the U.S Food and Drug Administration (FDA) issued a caution against chloroquine and hydroxychloroquine in the treatment of COVID-19 outside of hospital settings or clinical trials.[10] The FDA stated, “Hydroxychloroquine and chloroquine can cause abnormal heart rhythms such as QT interval prolongation and…ventricular tachycardia.” The FDA noted that QT prolongation was more common among persons receiving azithromycin and those with prior heart problems or kidney disease.

Hydroxychloroquine nonetheless has a 65-year track record of safety when prescribed at recommended doses in populations with normal liver and kidney function, and without pre-existing cardiac arrhythmias.[11] In the medical specialties of tropical medicine and rheumatology, chloroquine and hydroxychloroquine have routinely been prescribed without baseline laboratory testing or EKG monitoring. Whether these tests should be performed in the setting of hydroxychloroquine for COVID-19 is controversial. As it stands, a number of ongoing clinical trials have been paused or halted by regulatory authorities over concerns related to the potential for QT prolongation. Importantly, these safety concerns have risen from reports of hydroxychloroquine use in hospitalized patients, who are more likely to have severe infections, significant comorbidities and be on multiple concurrent medications.[12]

There are some features of the SARS-CoV-2 virus which may predispose individuals with COVID-19 to be more likely to have complications from drugs that prolong the QT than healthy individuals. SARS-CoV-2 itself can enter cardiomyocytes and may cause direct cardiac injury.[13-15] Multiple reports of increased arrhythmias in individuals with COVID-19 without other cause, suggest SARS-CoV-2 itself may cause arrhythmias.[15] Alternatively, elevated cytokines directly or in concert with cardiomyocyte damage, may predispose to arrhythmias.[16] Additionally, COVID-19 is associated with significant electrolyte imbalances, including sodium, potassium, and calcium as well as renal failure, each of which also predispose individuals to arrhythmias.[15, 17] Therefore testing the safety of hydroxychloroquine in individuals with COVID-19 specifically is valuable.

The safety of hydroxychloroquine use for COVID-19 in outpatients has not been established, but is believed to be less risky in outpatients than inpatients.[18] To address current knowledge gaps regarding the safety and tolerability of hydroxychloroquine in the outpatient prevention and treatment of COVID-19, and to inform its future usage in the setting of clinical trials, we present the safety data from three randomized placebo-controlled clinical trials of hydroxychloroquine in North America.

## Methods

### Study Design

We conducted three randomized, double-blind, placebo-controlled trials investigating hydroxychloroquine as prophylaxis and treatment for COVID-19 disease. The first two trials evaluated: 1) post-exposure prophylaxis (PEP); 2) preemptive early treatment (PET), (Clinicaltrials.gov Identifier: NCT04308668).[3, 19] Trial enrollment began on March 17, 2020, concluded on May 6, and follow up was completed on May 20, 2020. The third trial assessed pre-exposure prophylaxis (PREP) for COVID-19 (ClinicalTrials.gov Identifier: NCT04328467). Enrollment for this third trial began April 6 and ended May 26, 2020, with follow-up concluding on July 13, 2020.

In each of these trials, participants were randomized to receive placebo or hydroxychloroquine. The PEP trial required participants to have a known exposure to a lab-confirmed COVID-19 case within four days either as a household contact or as a healthcare worker or first responder. The PET trial enrolled persons with COVID-19 symptoms of four or fewer days duration and either lab-confirmed SARS-CoV-2 or high-risk exposure to a known case within 14 days of symptom onset. The PREP required persons to be high-risk healthcare workers or first responders with ongoing occupational exposure to COVID-19.

Hydroxychloroquine dosing for both the PEP and PET trials was 800mg load dosing, followed by 600mg in 6-8 hours, and then 600mg daily for five days in total. Participants were instructed to split their follow-up dosing in the event of gastrointestinal upset. In designing the trials, investigators chose doses within the existing FDA-approved dosing range that were modeled to achieve therapeutic concentrations from day 1 through 10.[20] Hydroxychloroquine dosing for PREP was dosed at 400 mg orally once, followed by 400mg 6 to 8 hours later, thereafter 400mg weekly or twice weekly for the duration of follow-up, up to 12 weeks. The placebo was dosed similarly.

### Study Participants

Participants were enrolled in the three trials via internet-based surveys throughout the United States and selected Canadian provinces. Full details are online (Clinicaltrials.gov Identifier: NCT04308668, NCT04328467).[3] Participants were excluded if they were <18 years old, had an allergy to hydroxychloroquine, retinal eye disease, known glucose-6 phosphate dehydrogenase (G6PD) deficiency, known chronic kidney disease, stage 4 or 5 or receiving dialysis, known porphyria, weight <40 kg, known QT prolongation, or receiving chemotherapy. Current use of hydroxychloroquine, azithromycin, or cardiac arrhythmia medicines (flecainide, amiodarone, digoxin, procainamide, propafenone, or sotalol) were also exclusion criteria. On April 20, 2020, the FDA required additional exclusions of structural or ischemic heart disease and personal or family history of cardiac QT prolongation, and medications that prolong the QTc interval.

Health Canada mandated additional exclusions for Canadian participants. Women who were pregnant or breastfeeding were excluded, as were patients with: severe diarrhea or vomiting; known cirrhosis with a history of encephalopathy or ascites; known prolonged cardiac QTc interval, history of ventricular arrhythmia or history of sudden cardiac death; patients taking additional medicines that had a high risk of prolonging the electrocardiogram QTc interval in conjunction with hydroxychloroquine. The Institutional Review Board (IRB) in Ontario also mandated that a physician perform a complete review of medications for participants above age 65, to exclude those with important drug-drug interactions.

Participants in the PEP and PET trials completed follow-up email surveys on Days 1, 3, 5, 10, and 14, whereas participants in the PREP trial completed weekly follow-up surveys. Surveys obtained self-report of study drug adherence, side effects, new COVID-19 symptoms, new COVID-19 testing, and hospitalization. For participants who stopped their study drug due to side effects or other reasons, we encouraged them to continue observational follow-up and completion of self-report surveys.

### Statistical Analysis

The analysis presented is primarily descriptive, summarizing the frequency of reported medication side effects and medication-related serious adverse events, such as hospitalization, life-threatening events, or deaths. With follow-up ongoing for the PREP study at the time of publication, data through June 25, 2020, are included in this analysis.

### Approvals

IRB approval occurred at McGill University, Clinical Trials Ontario, University of Manitoba, and University of Alberta, and the University of Minnesota.

## Results

A total of 2795 individuals were enrolled into the three trials. The combined median age was 40 years (interquartile range [IQR] 34-49) and 51% were women. The median weight was 79 kg (IQR 66-91). The majority of participants (74%) were healthcare workers and first responders. Approximately 66% of the participants were taking no chronic medications, and 59% had no chronic medical conditions. Demographic data are displayed in **Table 1**.

**Table 1.**
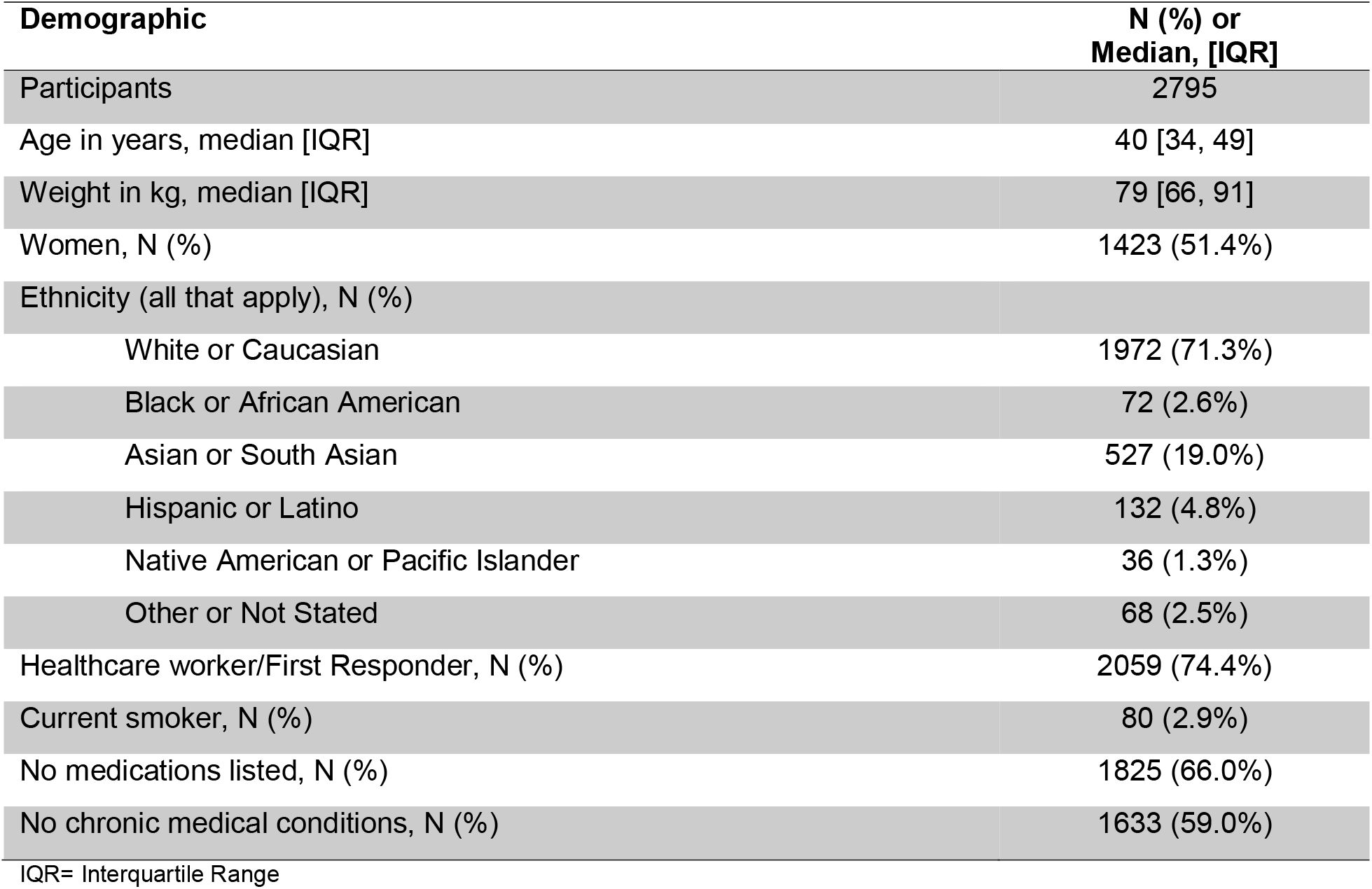
Baseline Demographics for all three cohorts, Post-exposure Prophylaxis, Preemptive Early Treatment, and Pre-exposure prophylaxis

### Post-exposure prophylaxis and early treatment trials

Of 1312 (n=821 PEP; n=491 PET) participants randomized, 87% (n=1139) started study drug and completed follow-up surveys. Of 130 participants who started drug but did not complete follow up, vital status was obtained for 32% (42/130), and all were alive. For the remainder, we performed an internet search for death records and found none.

Of 1139 who started study drug and reported side effect data, 29% reported one or more side effects, with more side effects reported among those on hydroxychloroquine (40%) versus placebo (18%) (**Table 2**). The most common side effects reported were upset stomach or nausea (25% on hydroxychloroquine versus 9% on placebo), followed by vomiting, diarrhea, or other GI symptoms (23% on hydroxychloroquine versus 6% on placebo), and neurologic reactions, such as lightheadedness or dizziness (7% on hydroxychloroquine versus 5% on placebo). Self-reported allergic reactions, occurred in 7 participants (6 on hydroxychloroquine and 1 on placebo). There were no reported episodes of arrhythmias or sudden cardiac death.

Only 46 participants (4%) from the PEP and PET trials reported that they stopped the 5- day treatment course due to side effects. The distribution of side effects was similar among both trials. No serious adverse events, resulting in hospitalization, attributable to medication side effects were reported.

**Table 2.** Combined Side Effects Days 1-5 in Post-Exposure Prophylaxis and Early Treatment Cohorts

|  | Hydroxychloroquine | Placebo | P-value* |
| --- | --- | --- | --- |
| Number randomized | 658 | 654 |  |
| Started study medication | 576 (87.5%) | 563 (86.1%) | 0.46 |
| Any side effect (days 1-5)** | 231 (40.1%) | 101 (17.9%) | **<0.001** |
| Side Effects** |  |  |  |
| Nausea or upset stomach | 146 (25.3%) | 53 (9.4%) | **<0.001** |
| Diarrhea, abdominal pain, vomiting | 131 (22.7%) | 35 (6.2%) | **<0.001** |
| Irritability, dizziness, vertigo | 39 (6.8%) | 26 (4.6%) | 0.13 |
| Tinnitus | 16 (2.8%) | 8 (1.4%) | 0.15 |
| Headache | 15 (2.6%) | 8 (1.4%) | 0.21 |
| Visual changes | 7 (1.2%) | 5 (0.9%) | 0.77 |
| Skin reaction | 10 (1.7%) | 4 (0.7%) | 0.18 |
| Taste change or dry mouth | 3 (0.5%) | 3 (0.5%) | >0.99 |
| Allergic reaction | 6 (1.0%) | 1 (0.2%) | 0.12 |
| Hot flashes, night sweats or palpitations | 2 (0.3%) | 1 (0.2%) | >0.99 |
| Fatigue | 1 (0.2%) | 1 (0.2%) | >0.99 |
| Panic | 0 (0.0%) | 1 (0.2%) | 0.49 |
| Other | 1 (0.2%) | 2 (0.4%) | 0.62 |

\*P values calculated via Fisher's Exact test. \*\*Of those who started study medication.

Since gastrointestinal issues are known to occur in COVID-19, we compared side effects between the placebo groups in the PEP group versus those who had COVID-19 in the PET group. We did not identify a statistical difference in the incidence of nausea / upset stomach reports in those with COVID-19 versus those exposed (11% vs. 8%, p=0.12). Similarly, the incidence of diarrhea, abdominal pain, vomiting, or other gastrointestinal issues did not differ between those with COVID-19 versus those exposed (7% vs. 4%, p=0.14). The risk of having any side effects did not differ by sex, age group, weight, or if one was a healthcare worker or not (**Table 3**).

**Table 3.**
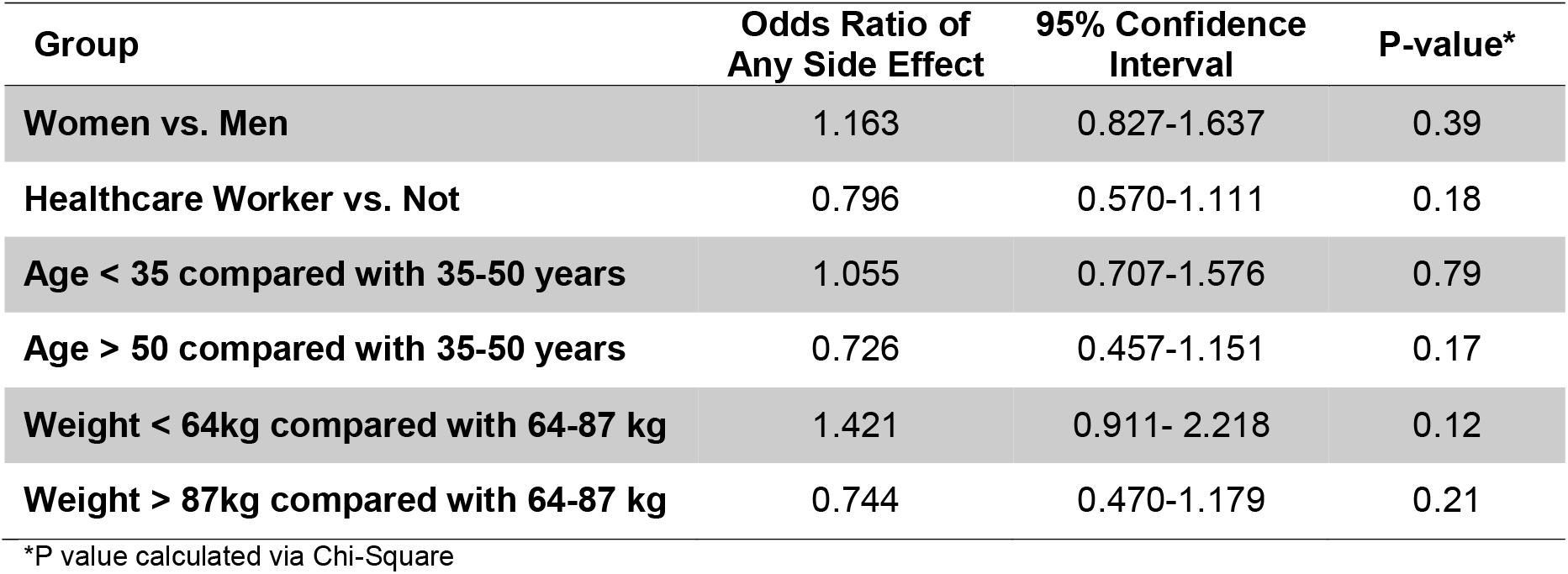
Odds of Side Effects in Post-exposure Prophylaxis and Early Treatment Cohorts.

### Pre-exposure prophylaxis trial

Interim data June 25, 2020, demonstrated that of 1483 randomized, 1402 had follow up data. Overall, 409 (29%) individuals experienced at least one side effect during the study (**Table 4**). Side effects were higher for those receiving hydroxychloroquine: 36% of those on twice-weekly dosing, 31% on weekly dosing, and 21% on placebo reported at least one side effect. Similar to the other trials, the most common side effects reported were upset stomach or nausea (18% on hydroxychloroquine twice weekly, 16% on hydroxychloroquine weekly, and 12% on placebo), followed by vomiting, diarrhea, or other GI symptoms (16% on hydroxychloroquine twice weekly, 12% on hydroxychloroquine weekly, and 6% on placebo).

**Table 4.** Side Effects from Pre-exposure Prophylaxis Cohort through June 25, 2020

|  | Hydroxychloroquine<br>2x per week | Hydroxychloroquine<br>1x per week | Placebo | P-value* |
| --- | --- | --- | --- | --- |
| Number randomized | 495 | 494 | 494 |  |
| Number with completed Surveys | 462 | 471 | 469 |  |
| Any side effect** | 167 (36.2%) | 145 (30.8%) | 97 (20.7%) | **<0.001** |
| Side Effects** |  |  |  |  |
| Nausea or upset stomach | 85 (18.4%) | 77 (16.3%) | 54 (11.5%) | **0.04** |
| Diarrhea, abdominal pain, vomiting | 75 (16.2%) | 56 (11.9%) | 30 (6.4%) | **<0.001** |
| Arrhythmia, tachycardia, palpitations | 8 (1.7%) | 8 (1.7%) | 11 (2.3%) | 0.73 |
| Irritability, dizziness, vertigo | 23 (5.0%) | 25 (5.3%) | 22 (4.7%) | 0.92 |
| Tinnitus | 6 (1.3%) | 10 (2.1%) | 4 (0.9%) | 0.26 |
| Fatigue | 4 (0.9%) | 0 (0.0%) | 1 (0.2%) | - |
| Visual changes | 4 (0.9%) | 7 (1.5%) | 3 (0.6%) | 0.41 |
| Skin reaction | 22 (4.8%) | 11 (2.3%) | 10 (2.1%) | **0.04** |
| Allergic reaction | 4 (0.9%) | 2 (0.4%) | 3 (0.6%) | 0.70 |
| Sleep disturbance | 6 (1.3%) | 8 (1.7%) | 4 (0.9%) | 0.52 |
| Myalgia | 1 (0.2%) | 4 (0.8%) | 2 (0.4%) | 0.38 |
| Arrhythmias | 1 (0.2%) | 0 (0.0%) | 1 (0.2%) | - |
| Other | 14 (3.0%) | 10 (2.1%) | 5 (1.1%) | 0.12 |

\*P values calculated via Fisher's Exact Test comparing hydroxychloroquine (pooled) versus placebo. \*\*Of participants who completed surveys

Two participants experienced atrial arrhythmias. One participant on placebo was hospitalized twice with atrial fibrillation. Another participant on hydroxychloroquine twice weekly was hospitalized after a syncopal event and was found to have a supraventricular tachycardia. No ventricular arrhythmias were reported.

In total, across the three cohorts, 25 individuals have been hospitalized through June 25, 2020 (1% incidence). The PEP trial had two hospitalizations, one in each arm and no deaths. The PET trial had 14 hospitalizations and two deaths. With hydroxychloroquine, 4 hospitalizations and 1 non-hospitalized death occurred. With placebo, 10 hospitalizations occurred with one hospitalized death. Two hospitalizations were for non-COVID-19, non-study medication related reasons, while the remaining 12 hospitalizations were due to COVID-19.

Nine individuals were hospitalized from the PREP study, one for COVID-19, two for arrhythmias (one on hydroxychloroquine twice a day and one on placebo), and the other seven hospitalizations were for non-study related reasons. No sudden unexplained deaths have been reported in any of the three trials. Full details are in **Supplemental Table 1**.

## Discussion

Among 2464 participants reporting on side effects from 3 randomized clinical trials investigating the efficacy of hydroxychloroquine in outpatient COVID-19 prevention and treatment, 27% reported at least one medication-related side effect. We captured one episode of supraventricular tachyarrhythmia with syncope in an individual on hydroxychloroquine and two episodes of atrial fibrillation in one individual on placebo. Whether hydroxychloroquine was responsible or contributed to the development of the supraventricular tachycardia is unclear. Hydroxychloroquine prolongs the QT interval and therefore is associated with an increased risk of ventricular arrhythmias, not atrial arrhythmias. Regardless, atrial arrhythmias were rare in both arms, and there were no episodes of ventricular arrhythmia or sudden death in approximately 50,000 patient-days of cumulative exposure.

As expected, the most common side effects were nausea and gastrointestinal upset. In addition to hydroxychloroquine, COVID-19 is also known to cause gastrointestinal upset. However, when we compared the rate of gastrointestinal upset among those receiving placebo in the post-exposure prophylaxis cohort versus those in the symptomatic cohort, reported gastrointestinal side effects were not significantly more frequent. Most participants regarded gastrointestinal upset upset as tolerable and completed the course of medication.

Medications causing QT prolongation are feared due to the risk of inducing ventricular arrhythmias, yet reports of arrhythmias due to hydroxychloroquine use are most often reported in the setting of co-ingestion, chronic use, or overdose.[21-23] The FDA does not recommend hydroxychloroquine be used with other agents that prolong the QTc,[11] such as azithromycin.[24] In 2017, the World Health Organization (WHO) reported that there has never been a reported sudden cardiac death attributable to chloroquine when prescribed at malaria treatment doses.[25] Despite the long history of chloroquine/hydroxychloroquine for malaria treatment and rheumatological diseases such as lupus, there have been increasing concerns around side effects in patients with COVID-19, especially related to arrhythmias.[26, 27] The FDA has now changed their emergency use authorization recommendation for hydroxychloroquine in COVID-19 treatment and prevention for this reason, among many others. [10]

For context, a perspective on dosing is needed to understand potential risks with chloroquine/hydroxychloroquine. Decades of safety data are available for the standard doses of chloroquine used for malaria prophylaxis (500 mg [300 mg chloroquine base] weekly) and malaria treatment (2.5 g [1.5g chloroquine base] total over three days). Cardiac conduction alterations were mostly seen when using much higher doses in hospitalized patients with COVID-19 (12 g chloroquine base over 10 days).[9] The cumulative dose of hydroxychloroquine-base used in our PEP and PET trials is 3.8 g (2.9 g base) total over 5 days. This is in line with established safe dosing strategies and below the doses shown to cause harmful effects.[28, 29] The Outcomes Related to COVID-19 Treated with Hydroxychloroquine among In-patients with Symptomatic Disease (ORCID) trial used 400 mg twice on day one followed by 200 mg twice daily for five days total (ClinicalTrials.gov Identifier: NCT04332991). They enrolled 479 individuals and also found no significant safety concerns at that dose.[30] Similarly, the RECOVERY trial used 2.4 g (1.86 g base) in four divided doses over 24 hours, followed by 800 mg (620 mg base) for an additional 9 days or until discharge and they reported no significant safety concerns in 11,000 hospitalized patients with COVID-19 randomized to hydroxychloroquine or placebo.[31]

Our trials additionally excluded participants taking azithromycin and other QT-prolonging drugs to enhance safety further. Other risk factors for QT prolongation, such as hypokalemia or hypomagnesemia, are uncommon in outpatients.[32, 33] Mercuro, et al. found QTc prolongation at 2.4 g courses of hydroxychloroquine over five days, but only clinically concerning side effects when combined with azithromycin and side effects were more common among those already on loop diuretics.[8] Our data showed one potential cardiac complication (atrial arrhythmia) in one individual on hydroxychloroquine and one on placebo. Our potential rate of atrial arrhythmias due to hydroxychloroquine is less than one in 1000. Many commonly used drugs, including antibacterial, antifungal, and other antimalarial drugs, are known to prolong the QT, thereby having a rare occurrence of arrhythmias.[34] The risk of QT prolongation has not precluded the use of drugs such as ciprofloxacin or fluconazole in most patients, but has required clinicians exercise caution.

Despite our rational design of these clinical trials, several limitations still exist. A significant limitation of our studies is related to their pragmatic nature, which relied on accurate self-reporting. We did not prospectively record or evaluate for laboratory abnormalities or QTc changes. While we have outcome data on 93% of participants who have completed the studies, it is possible, given the passive nature of self-reported follow-up, that those lost-to-follow-up may have had an adverse event that was not reported to our study team or their designated emergency contacts. Additionally, our cohorts are relatively young, with a median age of 40. They had few comorbidities and were on few medications. They were also predominantly healthcare workers, comprising a population of individuals with high health literacy. All these factors make our outpatient research participants healthier than most hospitalized patients in North America. Finally, our study exclusively included outpatients. Patients admitted to hospital are generally older and have more comorbidities. They also are more likely to have cardiac complications of COVID-19 and thus may be more susceptible to develop adverse effects from hydroxychloroquine.

## Conclusion

While efforts towards the development of a vaccine continue, agents that can prevent and treat COVID-19 are important. There is still equipoise concerning the potential efficacy of hydroxychloroquine as an effective agent against COVID-19; thus, timely completion of randomized placebo-controlled clinical trials is essential. Thus, large scale clinical trials to carefully evaluate the limited potential therapeutics that have been identified and perhaps even to replicate results of completed trials are imperative to our public health response. Ongoing clinical trials can safely continue with research participants and regulatory bodies reassured as to the general safety of hydroxychloroquine when using appropriate exclusion criteria.

## Data Availability

The data will be available in a public database 30 days after the pre-exposure prophylaxis trial is published.

## Acknowledgments

This work was supported by Jan and David Baszucki, Steve Kirsch, the Alliance of Minnesota Chinese Organizations, the Minnesota Chinese Chamber of Commerce, and the University of Minnesota. Personnel was supported through the Doris Duke Charitable Foundation through a grant supporting the Doris Duke International Clinical Research Fellows Program at the University of Minnesota. Katelyn Pastick and Elizabeth Okafor are Doris Duke International Clinical Research Fellows. Sarah Lofgren is supported by the National Institute of Mental Health (K23MH121220). Caleb Skipper is supported by the Fogarty International Center (D43TW009345). Drs. Melanie Nicol, Radha Rajasingham, and Matthew Pullen are supported by the National Institute of Allergy and Infectious Disease (K08AI134262, K23AI138851, T32AI055433). Margaret Axelrod is supported by NIH T32GM007347 and F30CA236157. Drs. Lee and McDonald receive salary support from the Fonds de recherche du Québec – Santé. Canadian funding was received from various sources. In Quebec, funds were received from the Clinical Practice Assessment Unit of the McGill University Health Centre and the McGill Interdisciplinary Initiative in Infection and Immunity’s Emergency COVID-19 Research Funding. In Manitoba, research support was provided from the Manitoba Medical Service Foundation. In Alberta, support was provided by Northern Alberta Clinical Trials and Research Centre. In Ontario, support was provided by the Research Institute of St. Joseph’s Hamilton, the St Joseph’s Hospital Foundation in London, and Bridge to Health Medical and Dental. Purolator Canada provided in-kind courier support for Canadian sites participating in the post-exposure and early treatment trials. Apotex Pharmaceuticals Canada provided a donation of some of the hydroxychloroquine tablets used. Rising Pharmaceutical donated some of the medicine for U.S. trials. COVID-19 Emergency Supplement funding was requested from the National Institutes of Health for each of the three trials.

## Clinicaltrials.gov Identifier

NCT04308668 for post-exposure prophylaxis and early treatment trials.

NCT04328467 for pre-exposure prophylaxis trial.

